# Effectiveness of Cluster-Level Behavioral Interventions to Reduce Salt Intake: A Systematic Review and Meta-analysis

**DOI:** 10.1101/2024.11.22.24317771

**Authors:** Sujiv Akkilagunta, Victoria Thomas, Kalaiselvi Selvaraj, Jaya Prasad Tripathy, Sitikantha Banerjee, Ranjan Solanki, Pradeep R Deshmukh

## Abstract

**Background:** The mean global salt intake is estimated at 10g/day much higher when compared to the WHO recommendation of less than 5g/day. Behavioral change interventions are most effective when applied at groups instead of individuals. The previous reviews on the topic did not compile evidence on cluster interventions.

**Objectives:** We conducted a systematic review and meta-analysis to find out the effectiveness of cluster-level dietary interventions in reducing average daily salt intake and mean 24-hr urinary sodium excretion among general population.

**Search Methods:** We searched the following research databases using search terms relevant to this review – PubMed, EmBASE, Web of Science, Global Index Medicus, LILACS, Cochrane CENTRAL, TRoPHI databases, CTRI and WHO-ICTRP.

**Selection criteria:** We included published randomized controlled trials and quasi-experimental studies with interventions for population groups that reflect the resident communities. There were no restrictions for age or gender. We excluded the studies targeting patient groups with specific underlying health conditions.

The primary outcomes were mean reduction in daily salt intake in g/day and the mean reduction in daily 24-hr Urine sodium output.

**Data collection and analysis:** Two authors independently screened the titles, abstracts and full-text articles. Two review authors independently extracted data and assessed the risk of bias. We classified the complex interventions into six categories based on the principles of health promotion. The duration of follow-up (outcome assessment after the intervention) was classified as short (≤ 6 months), medium (6 to < 12 months) or long-term(≥ 12 months). Risk of bias was assessed using RoB2 tool for cluster RCTs and ROBINS-I tool for Non-randomized studies of intervention. We pooled effect size estimates from individual studies using generic inverse variance method using a random-effects model.

**Main results:** We included 15 studies based on the selection criteria including 10 RCTs and 5 Non-randomised studies of Interventions (NRSI). Information and counselling measures, in the short-term, reduced the salt intake by 1.25g/day (95% CI: -1.9 to −0.6). At the medium-term, it was 0.47 g/day (95% CI: -0.81 to −0.14). On long term follow-up, it was 1.51g/day (95% CI: -2.62 to −0.4).

Use of salt-monitoring tools reduced salt intake by 2.48 g/day (95% CI: -4.66 to −0.3). Environmental modification measures did not reduce salt intake significantly.

**Discussion:** Information and counselling interventions in conjunction with other measures effectively reduced the salt intake. When quantified, there was an average reduction up to 1.5g/day. Salt monitoring tools showed a greater effect in reducing dietary salt.

A total of nine RCTs were assessed for risk of bias, of which five high-risk of bias. Among five Non-randomized studies of intervention, four of them showed serious risk of bias. An updated review in the future is likely to resolve these issues.

**Registration No.:** PROSPERO (registration ID: CRD42020168783)

## Introduction

Dietary salt reduction is an important precursor to reduce the global non-communicable disease (NCD) mortality. It is estimated that high salt intake contributes to 1.72 million cardiovascular deaths and 40.54 million DALYs (Disability-adjusted Life years) The Global action plan for NCDs targeted a 30% relative reduction in mean population salt intake by 2025.^1^ The benefits of salt reduction in reducing blood pressure and risk of coronary artery disease and stroke are well documented.^2,3,4^ The World Health Organization (WHO) recommends reduction in salt intake to less than 5g/day to reduce the disease risk. ^5^ Compared to other risk factors, the population exposure to high salt intake is more with an estimate suggesting that 45% of the global population is exposed.^6^ The global mean salt intake is estimated at 10 g/day and among Indians it is estimated at 10.98 g/day (95% CI 8.57-13.40).^7,8^ The global NCD target translates to a reduction of 3.3g of salt per day for Indians. However, to what extent the proven behavioral intervention strategies achieve reduction and the amount of reduction needs to be evaluated.

For behavioral risk factors, the population approach to prevention is expected to have a large overall effect when compared to the individual approach.^9^ Given this, multiple countries have initiated salt reduction programs. A Cochrane review by McLaren et al., summarized the effectiveness of these national programs on salt intake.^10^ But the review included uncontrolled program evaluation studies.

A similar review compiled studies on state-level and community level programs on reduction of salt intake. The majority of the studies included in this review were observational in nature.^11^ Community program evaluations are often uncontrolled and prone to self-selection bias. Non-randomized studies have a problem of attribution. Both these reviews have not quantified the pooled effect of the interventions. Hence, systematic review of controlled trials with pooled analysis is needed. Documentation of the interventions will also help us in understanding the type of interventions carried out and the feasibility of implementation.

We conducted a systematic review to determine the effectiveness of cluster-level dietary interventions in reducing average daily salt intake and mean 24-hr urinary sodium excretion among general population.

## Methods

### Search strategy and selection criteria

#### Types of studies

Randomized controlled trials with single or multiple arms and quasi-experimental studies, conducted globally and published from their start date to September 2021 were included in this search.

#### Types of participants

Studies with participants reflecting the general population were included. We did not place restrictions on gender, age and geographic area. Studies specifically conducted among patients or specific disease subgroups were excluded from the review.

#### Types of Interventions

We included cluster-based interventions aimed at behaviour change. Behaviour change may be obtained through health education given by a trained counsellor with or without the use of counselling aids . The following types of interventions were included - information campaigns, food product reformulation, pricing interventions, marketing restrictions and nutrition information on package. Since settings based interventions (workplace, school etc.,) reflect the general population, we included these in the review.

#### Outcomes

The outcome variables included were mean change in salt intake, mean reduction in daily sodium intake in (g/day) and mean change in 24-hr urinary sodium output.

#### Search Strategy

We searched the following research databases :PubMed, Embase, Web of Science, Global Index Medicus, Latin American Caribbean Health Sciences Literature (LILACS) database published by BIREME, Cochrane Central Register of Controlled Trials (CENTRAL), Trial Register of Promoting Health Interventions (TRoPHI) databases, Central Trials Registry-India (CTRI) and International Clinical Trial Registry Platform (ICTRP)-WHO-WHO-ICTRP.

This systematic review and meta-analysis was registered with PROSPERO (registration ID: CRD42020168783) and is designed, conducted and reported following the preferred reporting items for systematic reviews and meta-analyses (PRISMA) guidelines.

We developed a peer-reviewed detailed search strategy using the PICO principle by combining established search terms and free text terms for each database and trial registers. A search strategy for PubMed was initially developed which was adapted for the other databases and trial registers to account for differences in search terms and syntax rules. Two authors (SA and VT) jointly conducted the search using extensive keywords and Medical Subject Heading terms. The key search terms included: (“Sodium, dietary” OR “Sodium Chloride, Dietary”) AND (“substitut*” OR “reduc*”) AND (“program*” OR “strateg*” OR “initiative*” OR “Nutrition policy” OR “Food formulated” OR “Food industry” OR “Health Promotion” OR “Mass media”) (see Supplementary File S1 for full search strategy). Further, hand search was carried out by going through the references of the retrieved articles. We also performed related reference searches on PubMed to ensure that we had identified all available published material for each intervention.

#### Screening

The titles and abstracts of the reports identified through the online searching was screened for eligibility using a standard checklist. Full extract of all reports that appeared to meet the inclusion criteria, or for which titles and abstracts provided insufficient details were retrieved and reviewed by two independent researchers (SA and VT). The disagreements were resolved by discussion with other co-authors.

#### Data Extraction

Data were extracted from the studies meeting inclusion criteria by two independent researchers (SA and KS) using pre-tested data extraction forms. The data extraction form is attached as annexure in Supplementary File S1. Studies reported in other languages were translated before assessment. In case of multiple publications reported from a single research study, the record with the relevant data was included, however the data for methodology was extracted from other publications.

Risk of bias was assessed using the Revised Cochrane Risk-of-Bias tool (RoB2) for randomized controlled trials and ROBINS-I tool for quasi experimental studies, based on the domains provided in the tool. (Supplementary table-2) Two independent reviewers (SA and VT) assessed risk of bias (high, low, unclear) for each bias domain. Risk of bias assessment was based on information obtained from different sources (reports of the same study, information in trial registers).

#### Risk of Bias assessment

Risk of bias was assessed using RoB2 tool for cluster RCTs for the primary outcome -salt intake. Following domain was assessed in terms of low-risk, some concern and high risk: bias arising from the randomization process, bias arising from timing of recruitment of participants in a cluster RCT, bias due to deviations from intended interventions, bias due to missing outcome data, bias in measurement of the outcome and bias in selection of the reported result. Overall five RCTs were judged as high risk of bias since most of them showed some concern on two or more domains.^17,18,20,22^ We labelled three RCTs as having some concerns ^14,16,19^ and one RCT as having low-risk of bias^15^. The risk of bias graph and summary of all studies is presented in Fig. 4.1.1 and Fig. 4.1.2 respectively. The risk of bias for Non-randomized studies of intervention was assessed using ROBINS-I tool (results presented in Fig. 4.1.3.)

#### Data Synthesis

The data was compiled in MS Excel and entered into Review Manger to calculate the pooled treatment effects. The main outcome were Salt intake in g/day and 24-hr urine sodium output in mmol/L in 24-hr. We used mean differences (MDs) to estimate the pooled outcomes. We entered the data using the generic inverse variance method when the studies reported the effect estimates with their 95% CIs or standard errors. When the mean change with standard deviations for each group were reported, we entered the data using inverse variance method. When an analysis included studies that reported both types of outcomes, the final analysis was computed using the generic inverse variance method. For pre-post designs, the mean change (SE) was extracted instead of mean (SD) in each group to avoid inflating the sample size. When studies reported sodium intake instead of salt intake, it was converted into salt intake using the conversion factor of 2.542.^12^ When salt intake or sodium intake was reported per week or per month, it was converted to daily intake using the relevant conversion factor (7 or 30).

When the cluster studies adjusted for clustering effect in their analysis using multi-level modelling or other methods, these effect estimates were entered directly using the generic inverse variance method. When the studies did not adjust for clustering effect, we decided adjustment on a case-to-case basis. For cluster RCTs, we assumed a Intraclass correlation coefficient of 0.1 ^12^ , but we could not adjust clustering for pre-post designs or repeated cross-sectional designs. For missing data, such as standard deviations, we calculated these following the advice in the Cochrane Handbook. ^13^

#### Assessment of heterogeneity

The studies included in the review were described in the characteristics of studies table. We assessed homogeneity of the studies based on similarity of populations, interventions, type of outcomes, and follow-up times. We considered populations to be similar based on the age group two sub-groups - adults and children. The interventions were classified based on the activities employed in behavioural change. We regarded follow-up times of six months or less as short-term, between six months to less than a year as medium-term, and more than one year as long-term. We quantified the degree of heterogeneity using the I^2^ statistic, where an I^2^ value of 25% to 50% indicates a low degree of heterogeneity, 50% to 75% a moderate degree of heterogeneity, and more than 75% a high degree of heterogeneity. When moderate to high heterogeneity was identified, we explored it using subgroup analyses.

The following sub-group analyses were performed: primary salt-intake intervention or multiple risk-factor intervention, type of control – active or no-intervention control and type of setting – Worksite or family base intervention. We pooled effect size estimates from individual studies using Review Manager 5 (Review Manager 2014). Since the heterogeneity was moderate to high in most of the analyses, we used a random-effects model for pooling the effect estimates and the output was displayed using Forest plot. It was decided to assess funnel plot for assessment of reporting bias where ten or more studies were included in a meta-analysis.

## Results

We identified a total of 2,898 records in the initial search. After removing duplicates (n=514), a total of 2384 titles/abstracts were screened. Based on the screening, a total of 150 reports were sought for full-text scrutiny. Of these, we excluded 118 reports and included 32 reports (15 studies). (PRISMA study flow diagram, **Fig.1**). The range of sample sizes of included trials ranged from 68 (Takada, T et al 2016^13^), to 2566 (Li, N. et al^14^).

**Figure 1.**
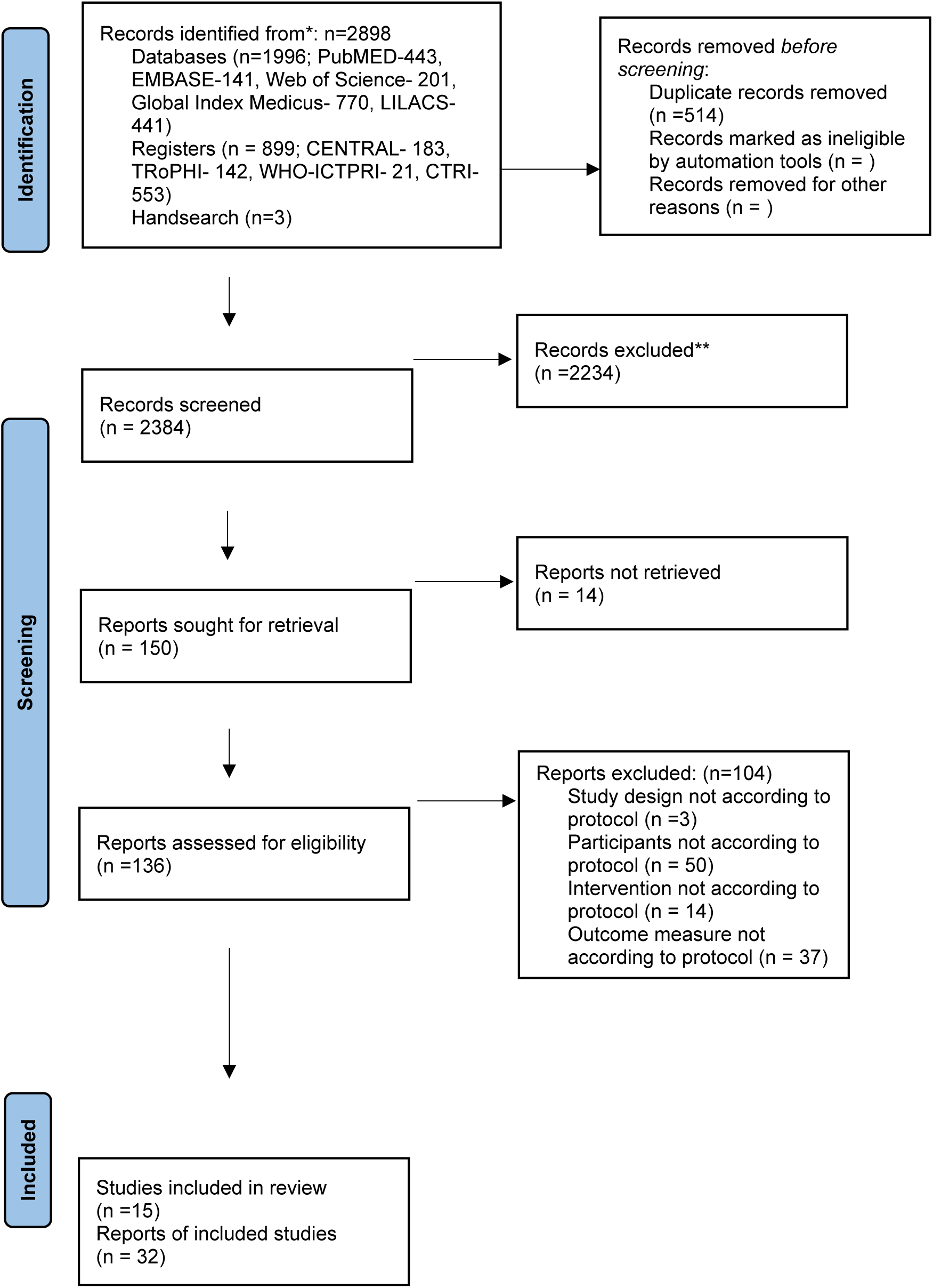
PRISMA study flow diagram

Out of the 15 included studies, 10 were cluster RCTs and 5 were Quasi-Experimental studies. All the RCTs had two arms except for that of Riis, N.L. et al^15^ which had three arms. Among the five quasi-experimental studies, two were controlled before-after studies (Geaney, F et al.; Jafari, M et al)^16,17^ , two were pre-post designs (Enkthungalag, M et al; Beer-Borst, S et al)^18,19^ and the other was a repeated cross-sectional design (Land, M.A. et al)^20^. The studies included in this review used multiple methods for achieving behavioural change. We grouped the interventions based on the means employed for behavioural change-Information & Counselling measures, Salt monitoring tools, Environmental modification, Provision of low-sodium salt substitute, Skill training in cooking low-sodium meals and Food product reformulation. Few studies used a single type of intervention. A brief overview of the studies included in the review on cluster-level studies have been provided in **Table-1**. The detailed description of the studies is presented in **Supplementary file S2**.

**Table 1.**
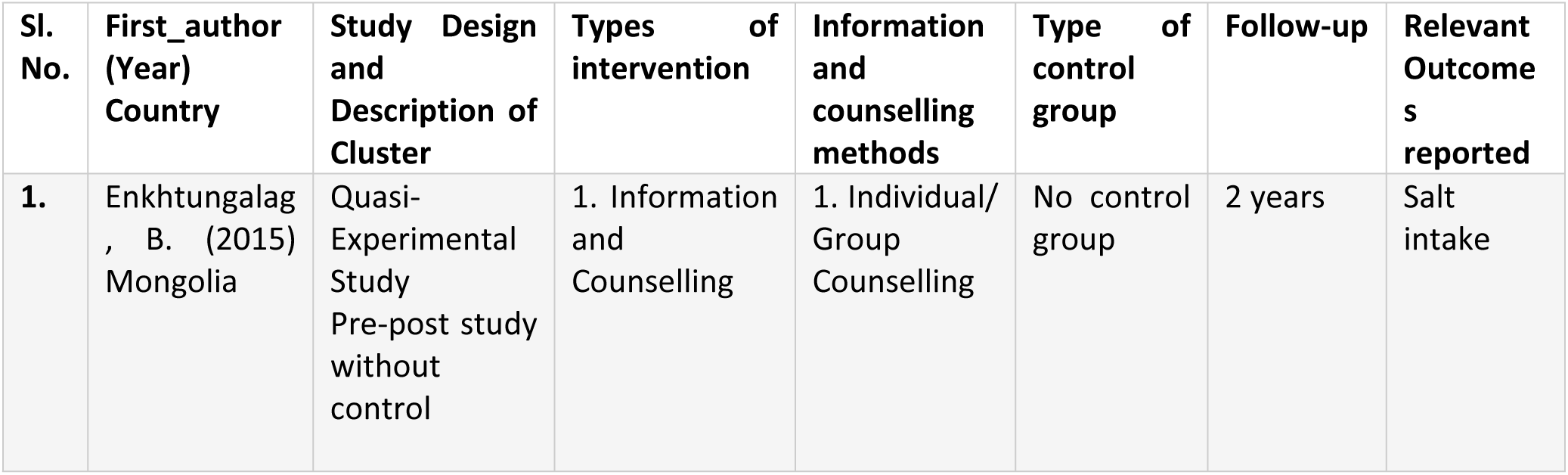
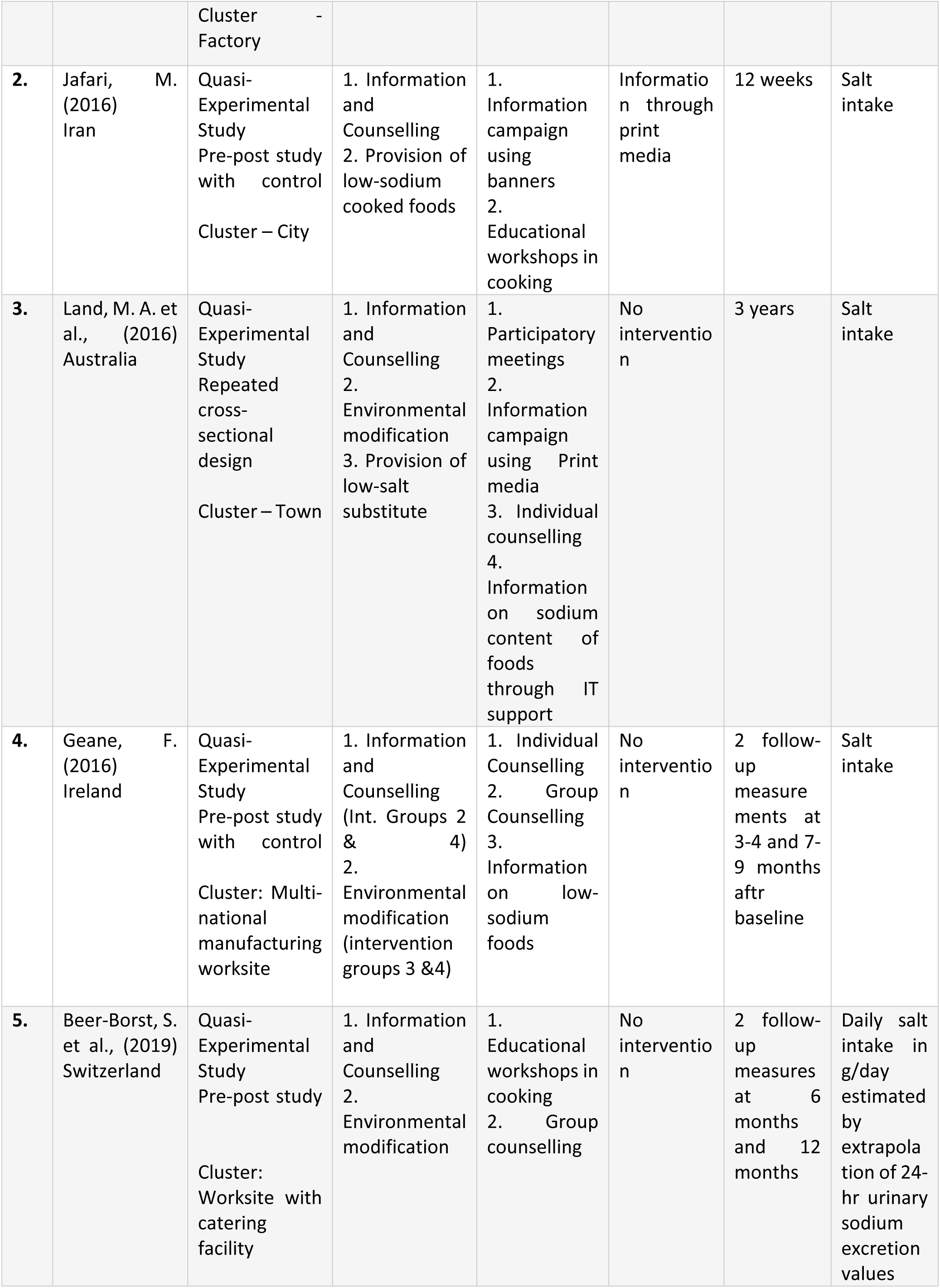
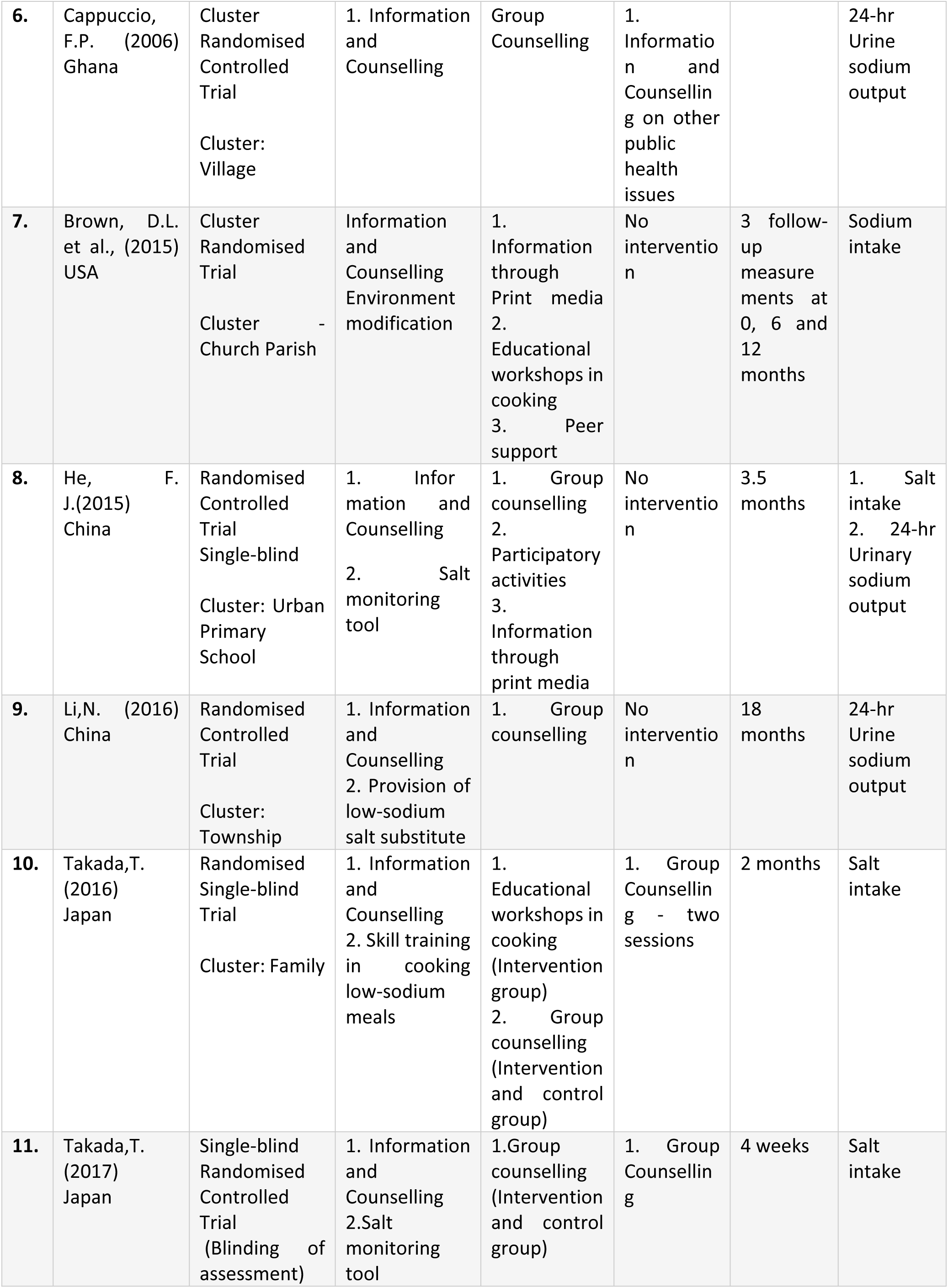
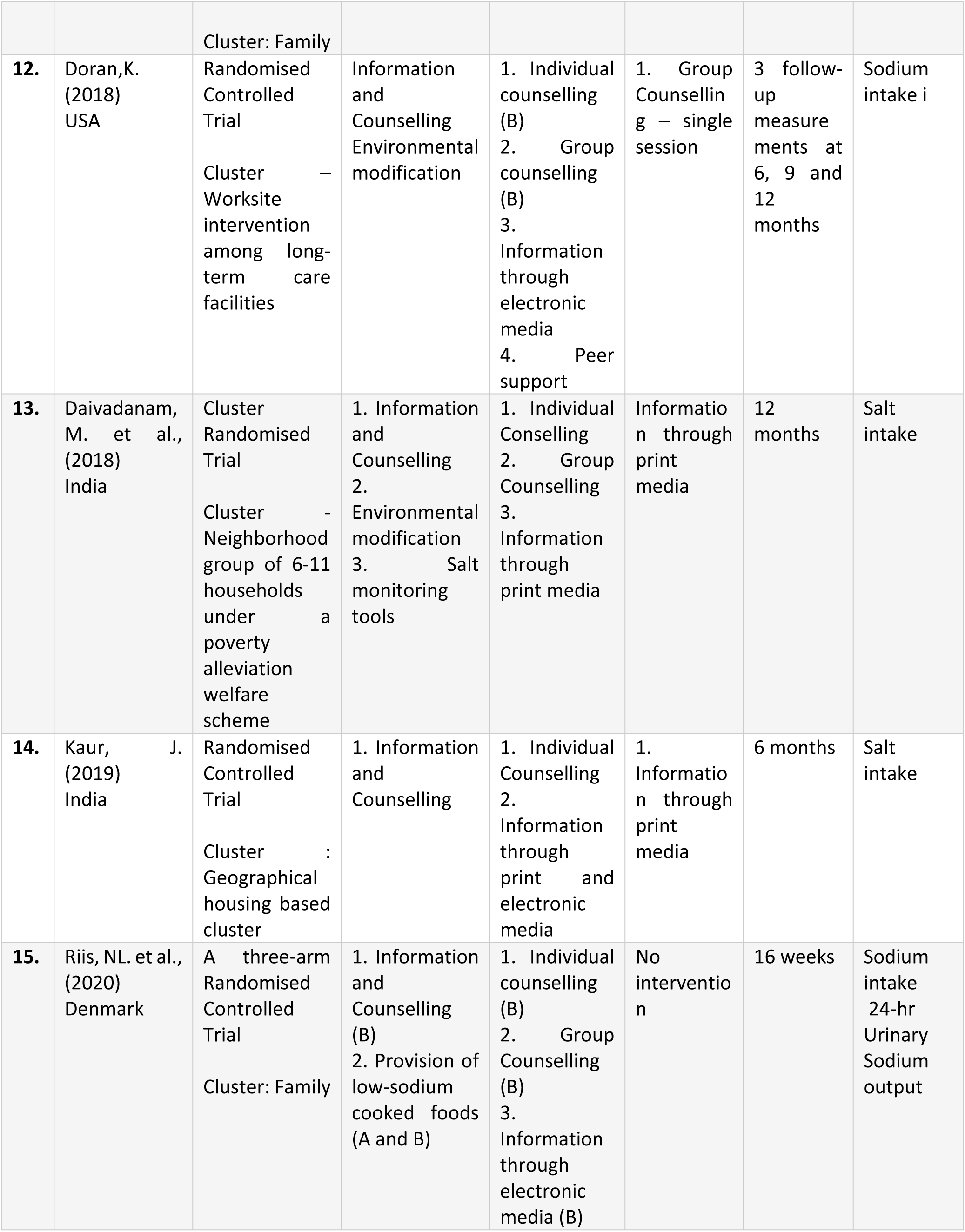
Brief Summary of the studies included in the review on cluster-level studies

### Risk of Bias assessment

We assessed risk of bias among the randomized trials using RoB2 tool for cluster RCTs for the primary outcome i.e., salt intake. Of the 10 RCTs, five were judged as high risk of bias.^17,18,20,22^ We labelled three RCTs as having some concerns ^14,16,19^ and one RCT as having low-risk of bias^15^. The risk of bias graph and summary of all studies is presented in **Fig. 2** and **Fig. 3** respectively.

**Figure 2.**
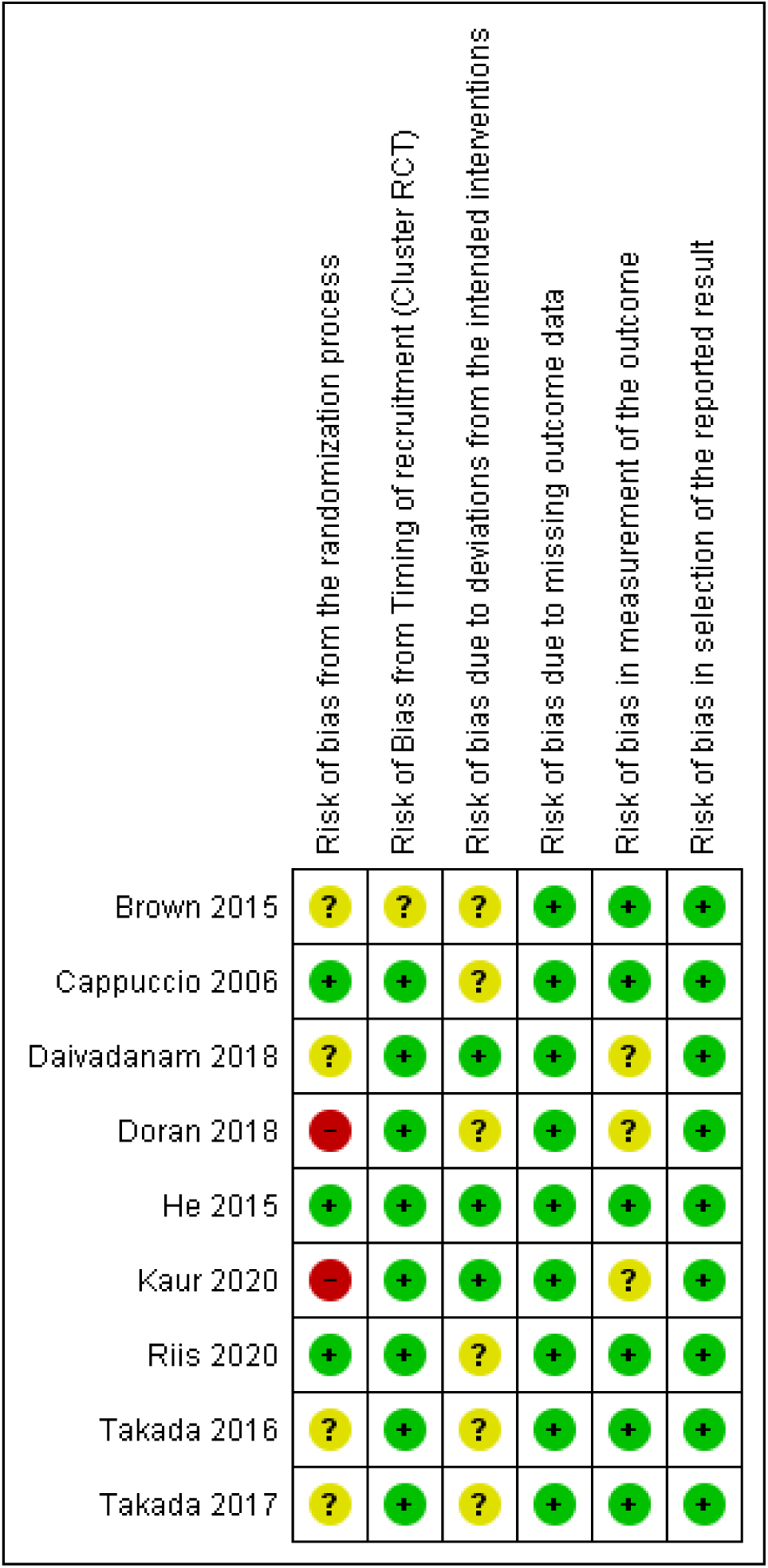
Risk of Bias (Summary) for the primary outcome (Salt intake) among the included cluster-RCTs using RoB2 tool

**Figure 3.**
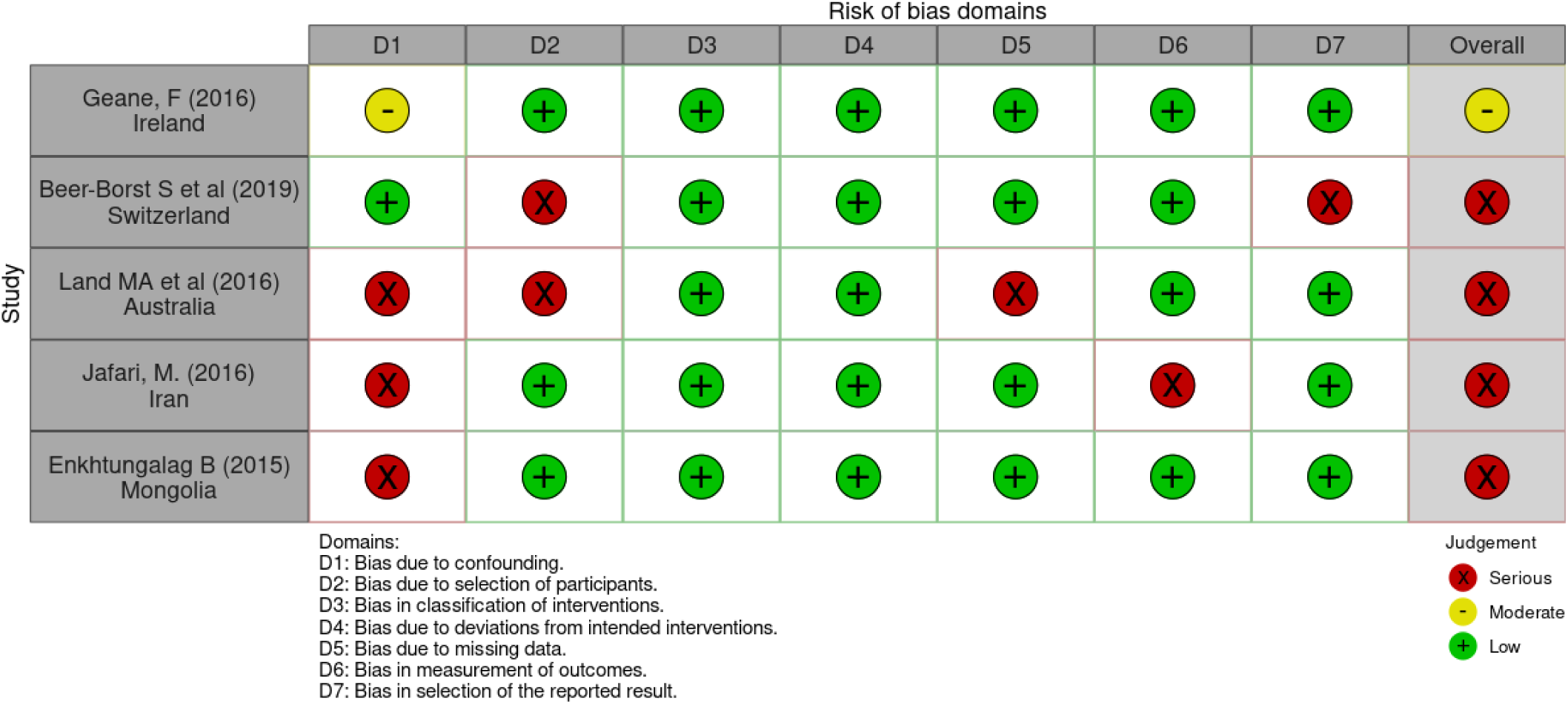
Risk of Bias for the primary outcome (Salt -intake) among the included NRSIs using ROBINS-I tool

### Effect of interventions with information and counselling measures on salt intake

Seven studies ^13,15,17,21–24^ evaluated the short term (<=6 months) effect of information and counselling measures on reduction of salt intake. Their meta-analysis revealed that the information and counselling measure alone or in combination with other interventions reduced the salt intake by 1.25g/day (95% CI: -1.9 to −0.6, I^2^ = 84%). Sub-group analysis revealed that RCT sub-group (6 studies) significantly reduced salt intake by 1.16g/ day (95% CI: -1.85 to −0.48, I^2^ = 85%) **(Fig. 4).**

**Figure 4.**
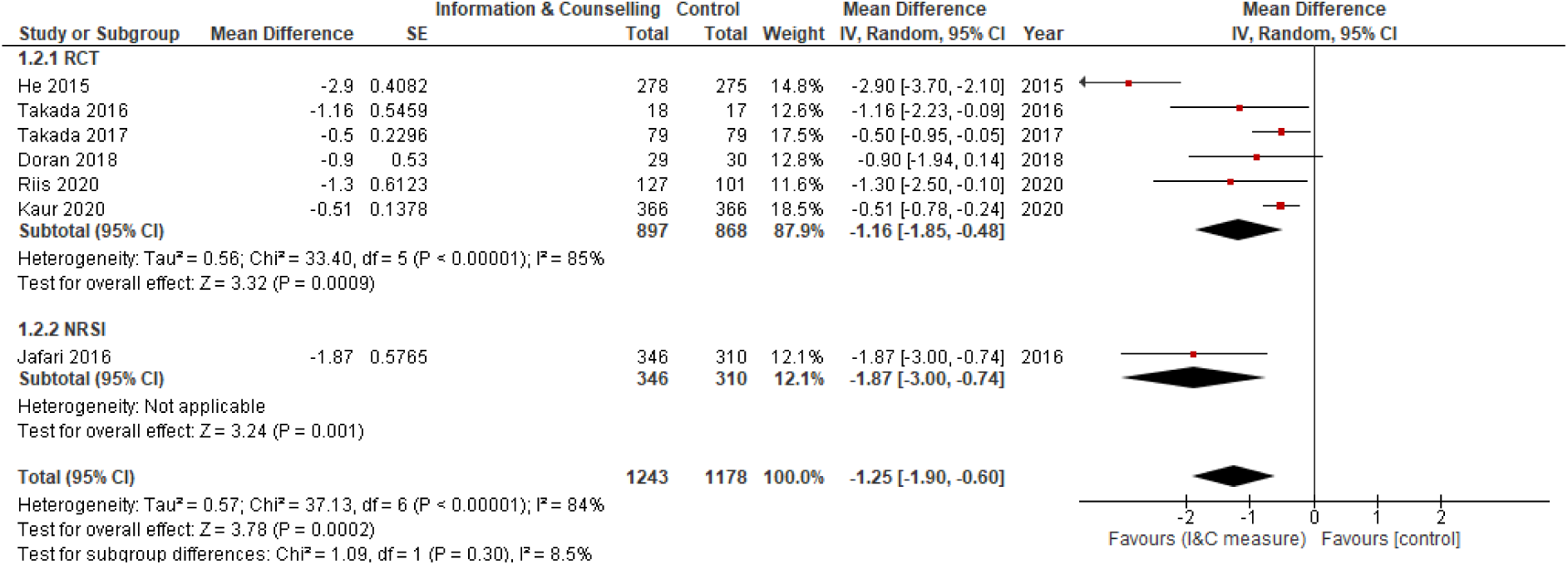
Forest plot showing effect of information and counselling measures on salt intake (g/day) in short term (<=6 months)

Two studies ^16,23^ evaluated the medium term (6 to <12 months) effect of the intervention (one RCT and one NRSI). Pooled analysis of the two studies revealed a reduction in salt intake by 0.47 g/day (95% CI: -0.81 to −0.14, I^2^ = 47%). (**Fig. 5**) We observed six studies^18–20,23,25,26^ reporting long-term effect (≥ 12 months) of information and counselling measures on salt-intake. Pooled analysis among them revealed an overall reduction in salt intake by 1.51g/day (95% CI: -2.62 to −0.4, I^2^ = 85%). But no significant reduction was noted in sub-group analysis (RCT and NRSI) (**Fig. 6**).

**Figure 5.**
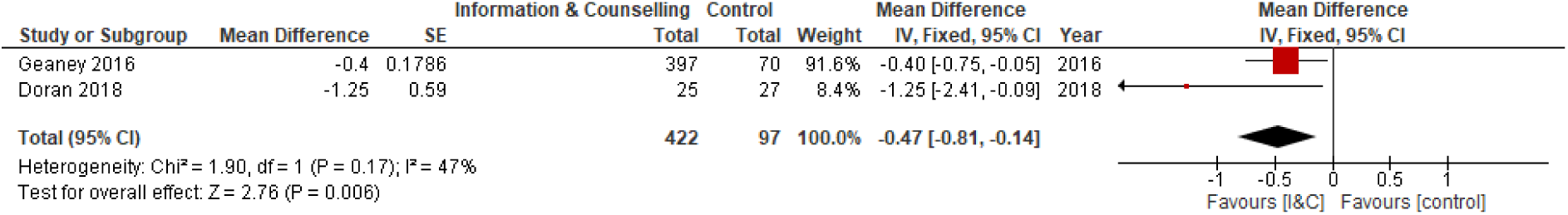
Forest plot showing effect of information and counselling measures on salt intake (g/day) in medium term (6 to <12 months)

**Figure 6.**
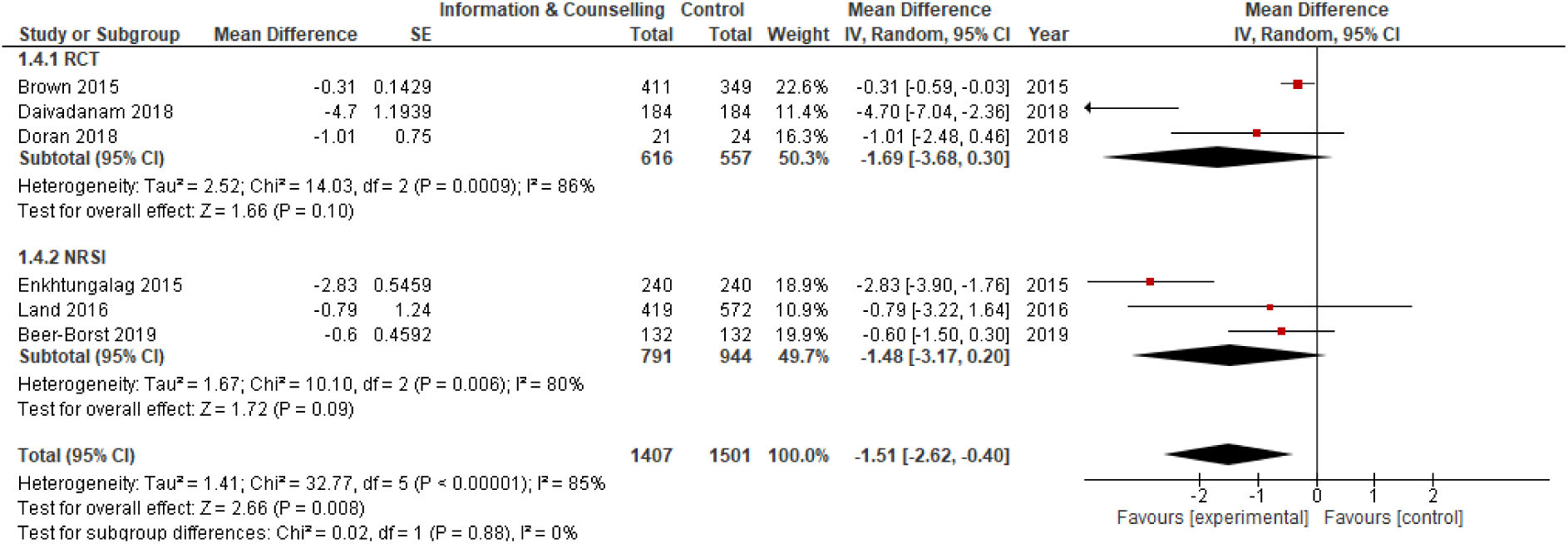
Forest plot showing effect of information and counselling measures on salt intake (g/day) in long-term (>=12 months)

### Effect of interventions with salt-monitoring tools in reducing dietary salt intake

Three studies^21,22,26^ reported the effect of salt-monitoring tools alone or in conjunction with other measures on salt-intake **(Fig. 7.)**. In the pooled analysis, there was a reduction in salt intake by 2.48 g/day (95% CI: -4.66 to −0.3, I^2^ = 94%). Sub-group analysis was not attempted due to few studies.

**Figure 7.**
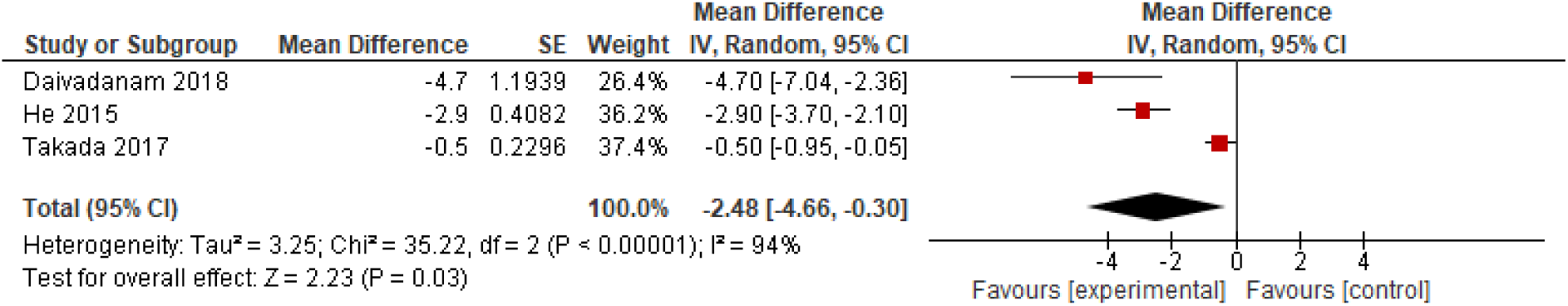
Forest plot showing effect of Salt monitoring tools on salt intake (g/day)

### Effect of interventions with environmental modification in reducing dietary salt intake

Five studies^16,18,20,25,26^ evaluated the effect of environmental modification measures alone or in conjunction with measures on salt-intake **(Fig. 8)**. In the pooled analysis, there was no significant reduction in salt intake (pooled mean reduction in 0.65 g/day [95% CI: -1.12 to −0.19, I^2^ = 75%]).

**Figure 8.**
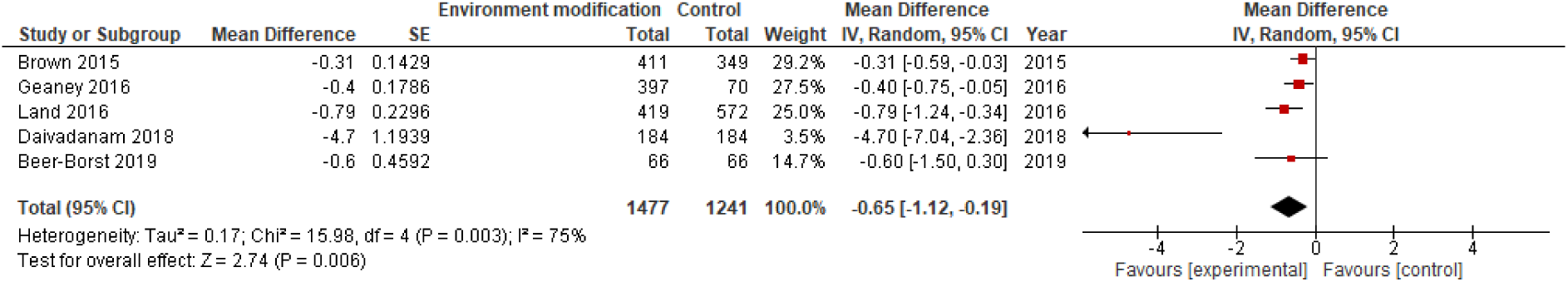
Forest plot showing effect of Environmental modification on salt intake (g/day)

### Effect of interventions with low-sodium cooked foods in reducing dietary salt intake

Two studies^15,17^ included provision of cooked foods alone or in conjunction with measures on salt-intake (**Fig. 6**.). In the pooled analysis, there was a significant reduction in salt intake (mean reduction was 1.60 g/day [95% CI: -2.42 to -0.78, I^2^ = 0%]).

### Effect of interventions at workplace settings in reducing dietary salt intake

Four studies including three NRSIs^16,18,19,23^ conducted in workplace settings with one or more measures for reduction in salt intake. In the pooled analysis **(Fig. 9)**, salt intake was significantly reduced by 1.15 g/day (95% CI: -2.21 to -0.1, I^2^ = 83%).

**Figure 9.**
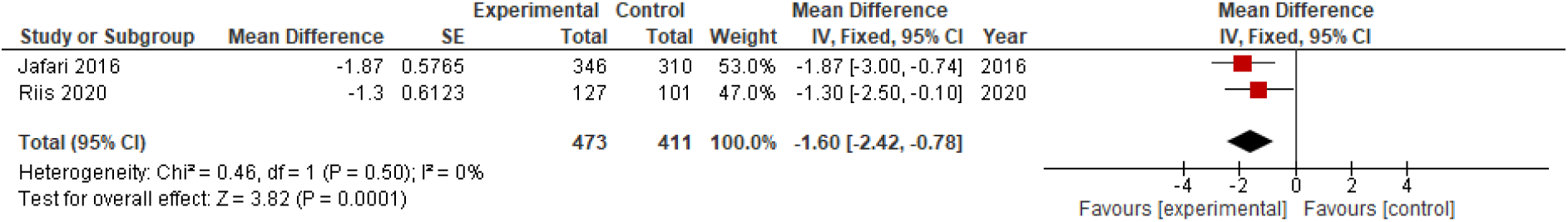
Forest plot showing effect of providing low-sodium cooked foods on salt intake (g/day)

### Effect of interventions at the household level in reducing dietary salt intake

Salt reduction by intervening among households was attempted in four RCTs. ^13,15,22,26^ In all studies, information and counselling measures were used as a part of the intervention. In the metaanlaysis **(Fig. 10)**, there was a significant reduction in salt intake – a reduction in 1.48 g/day (95% CI: -2.62 to - 0.34, I^2^ = 78%).

**Figure 10.**
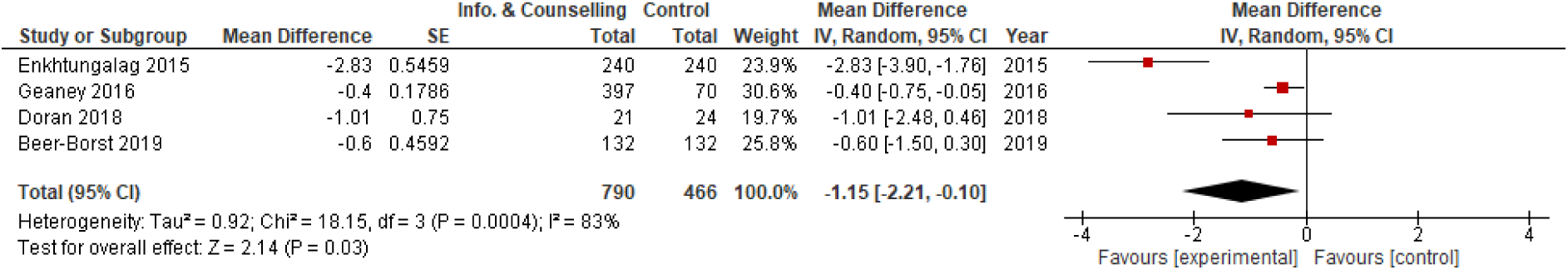
Forest plot showing effect of information and counselling measures given at workplaces on salt intake

The analysis for 24-hr urinary sodium output and comparison of unique salt-reduction interventions vs multiple risk-factor interventions is given in (**Supplementary file S2**). The effect of unique salt-reduction interventions was higher compared to when combined with other risk factors. We did not find any significant pooled reduction in 24 hr-Urine sodium output (**See Supplementary file S2**).

## DISCUSSION

We identified 15 cluster-intervention studies which evaluated interventions for reducing salt intake among general populations. Of these 5 were non-randomized studies of interventions. These interventions were categorized as – information and counselling measures, salt-monitoring tools, provision of low-sodium salt and low-sodium cooked foods.

Information and counselling measures were used in all studies alone or in conjunction with other measures. In the short-term (≤ 6 months), the interventions reduced the salt intake by 1.25g/day (95% CI: -1.9 to −0.6, I^2^ = 84%). At medium-term (6 to < 12 months), it was 0.47 g/day (95% CI: -0.81 to −0.14, I^2^ = 47%). In long term follow-up (≥ 12 months), there was an overall reduction in salt intake by 1.51g/day (95% CI: -2.62 to −0.4, I^2^ = 85%). The effect size was lower on medium-term follow up but only two studies supported this evidence.

Monitoring the salt intake could increase the awareness and thereby promote behavior change. Use of salt-monitoring tools as a part of the intervention reduced salt intake by 2.48 g/day (95% CI: -4.66 to −0.3, I^2^ = 94%). The heterogeneity among these studies was much higher when compared to other analyses. This could be due to difference in interventions– a spoon and a measuring kit were used by He et al^21^ and Daivadanam, M. et al^26^ respectively. The measurement of outcome also varied. Takada, T. et al., ^22^(2017) used an instrument for measuring 24-hr urinary sodium output as monitoring tool whereas Daivadanam, M. et al^26^ assessed self-reported outcome.

Environmental modification measures were those aimed at changing the socio-cultural and physical environment to enable the participants to consume low-sodium foods. Some of these interventions were made in workplace settings while others in the community. In work settings, the interventions included modification of canteen food by changing the menu and/or training the staff. In community the focus was to reduce accessibility and visibility of high-sodium preparations and vice-versa. When used in conjunction with other measures, environmental modification did not induce a large reduction in salt intake– with a moderate reduction of 0.65 g/day (95% CI: -1.12 to −0.19, I^2^ = 75%).

Pre-cooked or reformulated low-sodium foods were provided directly to the participants as a part of few interventions. However, there was no significant reduction in salt intake – a reduction in 1.12 g/day (95% CI: -2.28 to 0.05, I^2^ = 35%). Since there were only two cluster trials^15,17^, further studies are required for conclusive evidence. Reformulated low-sodium foods may help once a person is motivated for behavior change. Hence we propose such intervention should always be used in conjunction with other measures as is the case.

### Inferences and application of evidence

We included 15 cluster-based interventional studies to reduce salt intake among general populations. The studies were heterogenous in terms of the study setting, participants, intervention strategies, design and measurement of outcomes.

These studies were done in multiple strata of countries worldwide including high income countries (HICs) and low and middle income countries (LMICs). Hence the findings are applicable to most of the countries in the world.

All the studies used information and counselling measures as one of the intervention strategies. Information and counselling measures had significant impact on the reduction in salt intake in short-term. Dietary counselling is a simple means of intervention and feasible in most situations. But the intensity of counselling varied among all studies and it is difficult to generalize it as a uniform intervention.

Environmental modification measures, salt monitoring tools and provision of low-sodium reformulated foods also had significant impact when used in conjunction with other interventions. Of all these measures, salt monitoring tools had a higher effect and environmental modification measures had the least impact.

In both workplace and household settings the interventions showed significant impact, though the type of interventions and study design varied. We found that focused interventions solely aimed at reducing salt intake showed greater impact when compared to multiple risk factor interventions.(see file S1)

To the best of our knowledge, this is the only review that pooled the effectiveness of cluster-based salt-reduction strategies among otherwise healthy population sub-groups in terms of salt intake (g/day).

The average salt intake in India is estimated at 11g/ day.^27^ Since salt intake is socially influenced, population-based approaches have higher impact than individual-level approaches. The interventions at cluster level showed a modest reduction in salt intake ranging from 1.2 to 1.5g/ day. Salt monitoring tools used in conjunction with other measures showed a slightly higher average effect of 2.5 g/day. The description of interventions in this review can help in preparing and evaluating a nation-wide strategy to reduce salt intake.

### Quality of evidence

#### Risk of Bias in cluster RCTs

Of the 9 RCTs assessed for risk of bias, 5 of them had high-risk of bias. In most of these studies the risk of bias was due to randomization and allocation concealment process. Since these studies were cluster studies, allocation concealment was challenging. Many studies showed some concern in the domain – deviation from intended interventions. This was more often due to lack of adequate documentation rather than actual deviation from protocol.

#### Risk of bias in included studies – NRSIs

Among the Non-randomized studies of intervention most of the studies (4 of 5) showed serious risk of bias, though none of them had critical risk of bias. The risk of bias was mainly in two domains – confounding and selection of participants.

Since majority of the studies had high-risk (RCTs) or serious risk of bias (NRSIs), the quality of evidence in this systematic review may be considered as low to very low. However, the challenge of allocation concealment and blinding is persistent for conduct of high-quality cluster trials.

#### Potential biases, strengths and Limitations

We tried to pool the effect of salt-reduction interventions among general population. To the best of our knowledge, this is the first meta-analysis on dietary salt reduction interventions. We tried to include all the important databases to avoid any risk of publication bias. But since we did not include grey literature like conference abstracts, there was some possibility of excluding smaller studies. We tried to avoid language bias by searching regional scientific databases and not limiting to any language in the search strategy.

Generalizability may be limited due to wide heterogeneity among the studies. The attributability of a given intervention to the outcome was difficult due to multiple interventions. As newer studies are conducted in this area, these issues may be addressed by sub-group analysis and meta-regression.

We did not assess publication bias using a funnel plot owing to limitation in number of studies in each analysis (<10). Sub-group analyses and sensitivity analyses were done where feasible. Most sub-group analyses did not show an extensive variation in the effect estimate. The major variation was seen when single-risk factor and multiple risk factor studies were grouped separately. Such a variation in results is expected.

We included randomized and non-randomized studies in the meta-analysis, though non-randomized studies are considered inferior to randomized trials in terms of quality. This could be a major limitation in the study.

We pooled the outcome assessed by various methods in a single pooled analysis. This could have caused considerable heterogeneity.

#### Comparison with other studies

The present review focused on interventional studies that were conducted among otherwise healthy populations. We evaluated the primary outcome salt intake rather than blood pressure or other health related outcomes. There were similar reviews published in literature. Most of the reviews conducted were on hypertensive populations and evaluated blood pressure as the outcome.

Some reviews were restricted to regional population groups^28,29^. These reviews did not restrict to any population sub-group healthy or otherwise. For example, Jin, A. et al^28^ included studies that were conducted among hypertensive population, and healthy population. The reviews by McLaren, L. et al^10^ and Christoforou et al^11^ reviewed programmatic interventions. McLaren, L. et al^10^ reviewed programmatic interventions conducted among large population subgroups (country or state level) and obtained data from grey literature, in this case program reports. Most of them were not research studies but programs which had a pre-intervention and post-intervention data points. Due to wide heterogeneity in the population and interventions, they did not conduct a meta-analysis.

## Conclusion

Information and counselling interventions in conjunction with other measures effectively reduced salt intake. When quantified, there was an average reduction up to 1.5g/day. Salt monitoring tools showed a greater effect in reducing dietary salt.

## Data Availability

The search strategy used for various databases, the data extraction form are attached as supplementary files. The excel sheet used for metaanalysis will be available on request from the corresponding author.

## References

1. WHO. Global Action Plan for the Prevention and Control of Noncommunicable Diseases. World Health. Geneva; 2013.

2. Taylor RS, Ashton KE, Moxham T, Hooper L, Ebrahim S. Reduced dietary salt for the prevention of cardiovascular disease. In: Taylor RS, editor. Cochrane Database of Systematic Reviews. Chichester, UK: John Wiley & Sons, Ltd; 2011. p. CD009217.

3. WHO. Effect of reduced sodium intake on blood pressure, renal function, blood lipids and other potential adverse effects. Geneva; 2012.

4. Mozaffarian D, Fahimi S, Singh GM, Micha R, Khatibzadeh S, Engell RE, et al. Global Sodium Consumption and Death from Cardiovascular Causes. New England Journal of Medicine. 2014 Aug 14;371(7):624–34.

5. World Health Organization (WHO). Guideline: sodium intake for adults and children. Guideline: Potassium Intake for Adults and Children. 2012;1–46.

6. Murray CJL, Aravkin AY, Zheng P, Abbafati C, Abbas KM, Abbasi-Kangevari M, et al. Global burden of 87 risk factors in 204 countries and territories, 1990–2019: a systematic analysis for the Global Burden of Disease Study 2019. The Lancet [Internet]. 2020 Oct;396(10258):1223–49. Available from: https://linkinghub.elsevier.com/retrieve/pii/S0140673620307522

7. Johnson C, Praveen D, Pope A, Raj TS, Pillai RN, Land MA, et al. Mean population salt consumption in India. J Hypertens. 2017 Jan;35(1):3–9.

8. Adler AJ, Taylor F, Martin N, Gottlieb S, Taylor RS, Ebrahim S. Reduced dietary salt for the prevention of cardiovascular disease. Cochrane Database of Systematic Reviews. 2014 Dec 18;(12).

9. Rose G. Sick individuals and sick populations. Int J Epidemiol [Internet]. 2001 [cited 2022 Sep 2];30(3):427–32. Available from: https://pubmed.ncbi.nlm.nih.gov/11416056/

10. McLaren L, Sumar N, Barberio AM, Trieu K, Lorenzetti DL, Tarasuk V, et al. Population-level interventions in government jurisdictions for dietary sodium reduction. Cochrane Database of Systematic Reviews. 2016 Sep 16;(9).

11. Christoforou A, Trieu K, Land MA, Bolam B, Webster J. State-level and community-level salt reduction initiatives: a systematic review of global programmes and their impact. J Epidemiol Community Health (1978). 2016 Nov;70(11):1140–50.

12. Land MA, Webster J, Christoforou A, Praveen D, Jeffery P, Chalmers J, et al. Salt intake assessed by 24 h urinary sodium excretion in a random and opportunistic sample in Australia. BMJ Open [Internet]. 2014 [cited 2022 Sep 8];4(1). Available from: https://pubmed.ncbi.nlm.nih.gov/24440795/

13. Takada T, Imamoto M, Fukuma S, Yamamoto Y, Sasaki S, Uchida M, et al. Effect of cooking classes for housewives on salt reduction in family members: a cluster randomized controlled trial. Public Health. 2016 Nov;140:144–50.

14. Li N, Yan LL, Niu W, Yao C, Feng X, Zhang J, et al. The effects of a community-based sodium reduction program in rural China - A cluster-randomized trial. PLoS One. 2016;11(12):1–12.

15. Riis NL, Bjoernsbo KS, Toft U, Trolle E, Hyldig G, Hartley IE, et al. Impact of salt reduction interventions on salt taste sensitivity and liking, a cluster randomized controlled trial. Food Qual Prefer [Internet]. 2021;87(May 2020):104059. Available from: 10.1016/j.foodqual.2020.104059

16. Geaney F, Kelly C, Di Marrazzo JS, Harrington JM, Fitzgerald AP, Greiner BA, et al. The effect of complex workplace dietary interventions on employees’ dietary intakes, nutrition knowledge and health status: A cluster controlled trial. Prev Med (Baltim) [Internet]. 2016;89:76–83. Available from: 10.1016/j.ypmed.2016.05.005

17. Jafari M, Mohammadi M, Ghazizadeh H, Nakhaee N. Feasibility and Outcome of Reducing Salt in Bread: A Community Trial in Southern Iran. Glob J Health Sci. 2016;8(12):163.

18. Beer-Borst S, Hayoz S, Eisenblätter J, Jent S, Siegenthaler S, Strazzullo P, et al. RE-AIM evaluation of a one-year trial of a combined educational and environmental workplace intervention to lower salt intake in Switzerland. Prev Med Rep [Internet]. 2019;16(March):100982. Available from: 10.1016/j.pmedr.2019.100982

19. Enkhtungalag B, Batjargal J, Chimedsuren O, Tsogzolmaa B, Anderson CS, Webster J. Developing a national salt reduction strategy for Mongolia. Cardiovasc Diagn Ther [Internet]. 2015;5(3):229–37. Available from: http://www.pubmedcentral.nih.gov/articlerender.fcgi?artid=4451311&tool=pmcentrez&rendertype=abstract

20. Land MA, Wu JHY, Selwyn A, Crino M, Woodward M, Chalmers J, et al. Effects of a community-based salt reduction program in a regional Australian population. BMC Public Health [Internet]. 2016;16(1):1–11. Available from: 10.1186/s12889-016-3064-3

21. He FJ, Wu Y, Feng XX, Ma J, Ma Y, Wang H, et al. School based education programme to reduce salt intake in children and their families (School-EduSalt): cluster randomised controlled trial. BMJ. 2015 Mar 18;350(mar18 13):h770–h770.

22. Takada T, Imamoto M, Sasaki S, Azuma T, Miyashita J, Hayashi M, et al. Effects of self-monitoring of daily salt intake estimated by a simple electrical device for salt reduction: A cluster randomized trial article. Hypertension Research [Internet]. 2018;41(7):524–30. Available from: 10.1038/s41440-018-0046-0

23. Doran K, Resnick B, Alghzawi H, Zhu S. The worksite heart health improvement project’s impact on behavioral risk factors for cardiovascular disease in long-term care: A randomized control trial. Vol. 86, International Journal of Nursing Studies. Elsevier Ltd; 2018. 107–114 p.

24. Kaur J, Kaur M, Chakrapani V, Webster J, Santos JA, Kumar R. Effectiveness of information technology-enabled “SMART Eating” health promotion intervention: A cluster randomized controlled trial. PLoS One. 2020;15(1):1–22.

25. Brown DL, Conley KM, Sánchez BN, Resnicow K, Cowdery JE, Sais E, et al. A multicomponent behavioral intervention to reduce stroke risk factor behaviors: The stroke health and risk education cluster-randomized controlled trial. Stroke. 2015;46(10):2861–7.

26. Daivadanam M, Wahlström R, Sundari Ravindran TK, Sankara Sarma P, Sivasankaran S, Thankappan KR. Changing household dietary behaviours through community-based networks: A pragmatic cluster randomized controlled trial in rural Kerala, India. PLoS One. 2018;13(8):1–26.

27. Johnson C, Praveen D, Pope A, Raj TS, Pillai RN, Land MA, et al. Mean population salt consumption in India: a systematic review. J Hypertens [Internet]. 2017 [cited 2022 Sep 2];35(1):3–9. Available from: https://pubmed.ncbi.nlm.nih.gov/27755388/

28. Jin A, Xie W, Wu Y. Effect of salt reduction interventions in lowering blood pressure in Chinese populations: a systematic review and meta-analysis of randomised controlled trials. BMJ Open. 2020 Feb 17;10(2).

29. Tekle DY, Santos JA, Trieu K, Thout SR, Ndanuko R, Charlton K, et al. Monitoring and implementation of salt reduction initiatives in Africa: A systematic review. J Clin Hypertens. 2020;22(8):1355–70.

